# Biopsychosocial risk factors for Alzheimer’s disease and related dementias in UK immigrants from the Middle East and North Africa (MENA)

**DOI:** 10.64898/2026.05.08.26352762

**Authors:** Elizabeth Haddad, Ravi R. Bhatt, Akum Dhillon, Talia M. Nir, Lauren Salminen, Tala Al-Rousan, Kristine Ajrouch, Arpana Church, Nasim Sheikh-Bahaei, Neda Jahanshad

## Abstract

The Middle East and North Africa (MENA) region represents the area of greatest projected growth in instances of Alzheimer’s disease and related dementias (ADRDs) globally, yet, it remains virtually uncharacterized in health studies of aging and ADRDs. The UK Biobank is one of the largest and well characterized datasets of aging immigrants in the UK, offering an unprecedented opportunity to identify risk factors for ADRDs in individuals from MENA regions. Here we used the UK Biobank to compare sociodemographic, disease, lifestyle, genetic, and neuroimaging risk factors for ADRDs among UK immigrants from MENA countries (N=3,552) with two other large immigrant populations from Germany (N=1,097) and India (N=2,935), as well as a genetically British white control group born outside the UK (N=1,925). MENA immigrants exhibited a distinct and adverse risk profile characterized by greater socioeconomic deprivation, higher exposure to air pollution, poorer diet quality, lower physical activity, worse sleep, and higher smoking prevalence compared to European immigrant groups. The same trends were observed when comparing MENA to Indian immigrants, though these differences were less pronounced. These behavioral and environmental risk factors were accompanied by markedly higher rates of obesity, diabetes, hypertension, and other cardiometabolic conditions. Despite this substantial phenotypic burden, MENA participants carried a lower frequency of established AD genetic risk variants, including ApoE4, highlighting a discordance between genetic risk and observed disease related vulnerability. Neuroimaging analyses revealed lower hippocampal volume in MENA and Indian participants relative to European groups despite younger average age, consistent with early limbic vulnerability associated with metabolic and inflammatory stress. Overall, our results indicate that dementia risk in MENA populations is driven by a multidimensional framework of metabolic, systemic, and social environmental exposures that may shape vulnerability independently of canonical European-derived genetic risk factors. These findings highlight the urgent need for ancestry- and context-specific frameworks to support equitable dementia prevention and avoid under-predicting risk in underrepresented populations.

## Introduction

Understanding aging pathophysiology and risk for disease in diverse ethnoracial backgrounds is critical to provide effective and optimized healthcare to all populations. The Global Burden of Disease Study (GBD) 2019 estimated a 166% global increase in Alzheimer’s disease and related dementias (ADRD) cases by 2050. While age-standardized prevalence rates for dementia are stabilizing in many high-income countries, the Middle East and North Africa (MENA) region faces a disproportionate surge. By 2050, the MENA region is projected to see a 367% growth in ADRD prevalence, the highest increase globally (Nichols et al., 2022). Notably, recent systematic analyses found that the MENA region already has the highest age-standardized DALY rates (disability-adjusted life-years) for dementia among all global “super regions^1^” (GBD 2019 North Africa and the Middle East Neurology Collaborators, 2024). Yet, people from MENA countries are highly underrepresented in health studies of aging and ADRDs. For example, individuals of MENA ancestry have been left out of almost all genome-wide association studies (GWAS) of human traits, despite making up nearly 10% of the global population. These disparities contribute to the inability of largely European GWAS results to replicate across ethnic groups, and not only hinders translation to clinical practice, but could dangerously influence public health policy (Sirugo et al., 2019). Given the complex and polygenic nature of late onset dementias (typically occurring after age 65), addressing the current disparities in health studies of ADRD are paramount to change the current prevalence projections of ADRDs and future therapeutic planning.

Much of the projected surge in ADRD cases in MENA reflects rapid population growth and aging, which are the primary drivers of these forecasts. This demographic shift represents a relatively recent transition and the specific biological and cognitive pathways of decline in these populations are not yet fully understood. While demographics drive the absolute number of cases, recent systematic analysis showed that the rate of dementia-related DALYs that is potentially preventable by addressing modifiable risk factors is 39% higher in the MENA region than the global average (154.7 vs. 111.3 per 100,000 people) (GBD 2019 North Africa and the Middle East Neurology Collaborators, 2024). Specifically, metabolic risk factors, including high BMI and high fasting plasma glucose, and behavioral factors such as tobacco use are the primary modifiable drivers of dementia-related disability in this region. ADRD risk is also influenced by sex, with this disparity appearing more pronounced in MENA populations. In several MENA countries, obesity is twice as prevalent in women than in men (NCD Risk Factor Collaboration (NCD-RisC), 2017). Obesity, which is strongly shaped by environmental factors such as diet and stress, has been linked to alterations in brain structure and function, as well as an increased risk of ADRD (Arnoldussen et al., 2014). While obesity and related lifestyle factors are largely modifiable, they are also complex, influenced by the interplay of sociocultural, environmental and genetic factors. Therefore, understanding differences in risk profiles across ethnic populations is essential for developing a more precise assessment of the bio-social-environmental contributors to ADRD risk in MENA populations.

Importantly, much of an individual’s risk based on these factors would not decline if the person migrated to a lower-risk country. Over the last five decades, Western Europe and North America regions have seen a growing influx of immigrants, refugees, and asylum seekers from MENA countries. Many of these immigrants are motivated to do so due to political instability in their countries of origin and to obtain economic opportunities. The MENA population experiences significant health disparities and barriers to their unmet health needs, similar to the levels experienced by African and Hispanic immigrants (Ajrouch et al., 2024; Kindratt et al., 2023; Patel et al., 2022). Yet there is limited knowledge about how environmental stressors and sociocultural factors (e.g., immigration circumstances, language barriers, social isolation, xeno- and Islamophobia, discrimination, challenges acquiring secure employment and housing) interact with biology to increase risk for negative health outcomes and ADRD (Patel et al., 2022). Though as of yet there are no data that indicate prevalence of ADRD in MENA immigrant populations, a growing body of literature in the U.S. illustrates that MENA immigrant adults report higher rates of cognitive impairment (Al-Rousan et al., 2023; Dallo et al., 2021; Kindratt et al., 2022; Kindratt & Smith, 2024) and worse overall cognitive health (Ajrouch et al., 2024; Zahodne et al., 2023) compared to White and Black adults. Large biobank data sets may be an untapped resource to begin to identify mechanisms driving cognitive health disparities and their biological underpinnings.

Global strategies for dementia prevention are increasingly guided by the Lancet Commission’s framework, which in its 2024 update identifies 14 modifiable risk factors across the life course that offer a critical pathway for reducing dementia risk (Livingston et al., 2024). The UK Biobank (UKB) is a large, densely phenotyped study on aging that provides a unique opportunity to operationalize the Lancet Commission’s framework of modifiable risk factors for ADRD among underrepresented immigrant groups in the United Kingdom (Bycroft et al., 2018). Its rich characterization of sociodemographic, disease, lifestyle, and genetic variables enables a systematic assessment of how life-course modifiable factors contribute to distinct ADRD vulnerability observed in MENA populations. Importantly, the UKB also includes multimodal brain MRI, offering a non-invasive window into early structural and microvascular changes associated with brain aging and ADRD risk - an area of science that has been grossly understudied among MENA populations.

The purpose of this study is to characterize sociodemographic, disease, lifestyle, genetic, and neuroimaging risk factors for ADRDs in UK immigrants from MENA countries using the UKB dataset. While the Lancet Commission’s framework of 14 modifiable risk factors provides a critical baseline, our analysis extends to a broader array of GBD-defined risk factors. We compared these factors to those of other immigrant populations from India (the largest immigrant group in the UKB), and Germany (the largest European immigrant group), as well as a genetically British-White control group born outside the UK in either MENA countries, India, or Germany. By integrating biological, lifestyle, and environmental determinants of brain aging, our goal is to provide the first population-level description of dementia-related risk architecture in MENA individuals and to identify modifiable targets for equitable dementia prevention.

## Methods

### Study population

The UKB is a large population-based study that has collected deep phenotypic and genetic data from approximately 500,000 community-dwelling adults across the United Kingdom. Participants, aged 40–69 years at the time of recruitment, were identified via the National Health Service (NHS) register and invited to volunteer for the study, provided they resided within a reasonable traveling distance of a dedicated assessment center. Recruitment and the baseline assessment visits took place between 2006 and 2010 at multiple assessment centers across the UK. During the visit, participants provided electronic signed consent, answered touchscreen questions on health, sociodemographic and lifestyle factors, and completed a range of physical measures and cognitive assessments, while also providing biological samples (blood, urine, and saliva) for genetic testing and further assay. A subset of these participants were subsequently invited back for further assessment, including a multi-modal brain imaging scan that began to take place in 2014. (Bycroft et al., 2018). Participant selection is detailed in **Figure 1**. Data from subjects with genotyping (n∼500k) were initially screened to determine individuals born outside of the UK (n=40,571). Inclusion criteria for MENA populations were based on the union of 7 (World Bank (World Bank, 2003), FAO (Santos & Ceccacci, 2015), UNAIDS (Dungerdorj et al., 2019), UNICEF (UNICEF Middle East and North Africa Region, 2024), UNHCR (van Waas, n.d.), IMF (Davoodi & Abed, 2003), and UNSD (United Nations Statistics Division, 1999)) definitions of the Greater Middle East or Middle East and North Africa countries. A complete list of counts and countries included may be found in **Table S1**. While our primary definition of MENA is intentionally broad, we also conducted sensitivity analyses using the GBD definition of the North Africa and Middle East “super-region”; these results are detailed in the supplementary materials (**Tables S51–S73**). Comparison groups included immigrants born in India (self-identified as Indian; the UK Biobank’s largest immigrant group) and immigrants born in Germany (the largest European immigrant group) as a control group of European ancestry. A third comparison group included individuals born in any of the above mentioned countries having genetically determined British-White ancestry (British; non-UK born) to account for some degree of variance associated with being an immigrant. Genetic, disease, and lifestyle variations were assessed across all immigrant populations (N=9509) at their baseline visit (2006-2010). Individuals with brain imaging available provided a subset (total N=626; MENA N=157; British N=212; Germany N=109; India N=148) in which imaging-derived phenotypes could be assessed.

**Figure 1.**
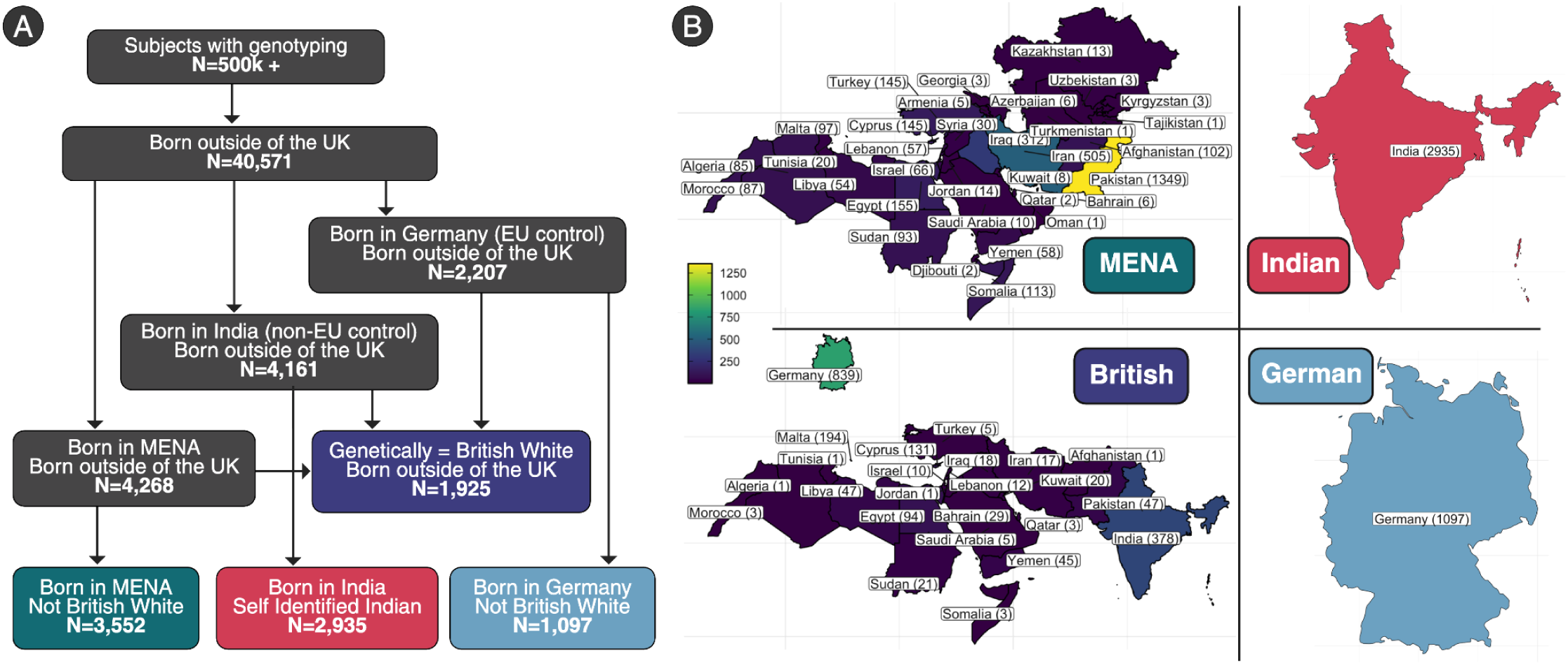
Participant selection. **A.** Workflow of participant selection. Immigrant participants with genotyping were parsed into 4 groups: those born in MENA, India, and Germany along with a genetically British immigrant control group comprising individuals born in any of the aforementioned countries. **B.** Color coded world maps for each group indicated the number of participants from each birth country.

### Demographic and lifestyle assessment

Demographic factors of interest assessed as part of the UKB study included age at first assessment, age and year of immigration to the UK, sex, college attendance (binary), and Townsend deprivation index (TDI). Environmental and pollution measures were also included. Lifestyle domains included variables related to diet, supplements, anthropometric measures, sleep, physical activity, social contact, blood pressure, smoking status (coded as “never”, “previous”, and “current”), and alcohol intake frequency (coded as “infrequent”, “occasional”, and “frequent”). From the data provided by UKB, summary/composite scores for diet, sleep, and social isolation/loneliness were computed for each category based on prior work ((Said et al., 2018; Zhuang et al., 2021), (Fan et al., 2020), and (Elovainio et al., 2017) respectively). Briefly, the overall diet quality score was computed as the sum of individual diet components that corresponded to ideal (score of 10) and poor (score of 0) consumption of various diet components. Ideal for fruit, vegetable, whole grains, fish, dairy, and vegetable oil generally meant a higher consumption was better, whereas ideal for refined grains, processed meat, unprocessed meat, and sugary food/drink intake meant lower consumption was better. Sleep scores ranged from 0 to 5 with higher scores indicating a healthier sleep pattern. Overall sleep patterns were defined as “healthy” (score ≥4), “intermediate” (score 2-3), and “poor” (score ≤1). Individuals were considered socially isolated if they scored 2-3 on a socially isolation score ranging from 0-3 and loneliness classification was determined if an individual scored 2 on a 0-2 loneliness scale. Prevalence of disease diagnoses were also calculated and visually displayed across the 4 ethnic populations in 5 year age bins. These included dementia, sleep apnea, stroke, heart failure, anxiety, atrial fibrillation, depression, cancer, coronary artery disease, headaches and migraines, diabetes, obesity, hypercholesterolemia, and hypertension.

### Imaging biomarker assessment

We extracted imaging measures commonly associated with aging, neurodegeneration, and cerebral small vessel disease. Structural T1w measures included average cortical thickness and hippocampal volume derived from FreeSurfer version 7.1.1. Diffusion weighted imaging measures included diffusion tensor imaging (DTI) metrics such as global fractional anisotropy (FA) and mean diffusivity (MD) derived from the ENIGMA TBSS pipeline (Kochunov et al., 2022). The global measure was calculated by weighing the skeletonized measures within each JHU region of interest (ROI) by the total number of voxels within that region and taking the average of all weighted measures across all ROIs. The presence of cerebral microbleeds (CMBs) were assessed using susceptibility weighted imaging following the Microbleed Anatomical Rating Scale (MARS) criteria (Gregoire et al., 2009), where our classifications were validated using other sequences to exclude CMB ‘mimics’. The number and position of CMBs were noted as well as their classification including ‘definite’, ‘possible’ and ‘total’ (definite + possible). Results here are reported for ‘total’ CMB counts. White matter hyperintensities (WMH) were assessed using a FLAIR (Fluid Attenuated Inversion Recovery) sequence and quantified using the BIANCA segmentation tool, as provided by the UKB (Griffanti et al., 2016).

### Genetic analysis and family history

We analyzed genotype data from UKB released in May 2018. The data was collected from 489,212 individuals, of which 488,377 passed quality control checks by UKB. Genotypes were then imputed using the Haplotype Reference Consortium (HRC) reference panel and a combined reference panel of the UK10K and 1000 Genomes projects Phase 3 (1000G) panels (Bycroft et al., 2018). There were 16,043,095 single nucleotide polymorphisms (SNPs) following quality control (QC),which included having a genotyping call rate (SNPs missing in individuals) >95%, removing variants with a minor allele frequency <0.01 (1%), removing variants with Hardy-Weinberg equilibrium p-values <1110^-6^, and removing individuals with greater than three standard deviations from the mean heterozygosity rate. All genomic processing was completed using *plink (Purcell et al., 2007)*. To develop a reference panel including MENA individuals from the UKB, genotypes from the HapMap3 (International HapMap 3 Consortium et al., 2010) study were combined with 137 whole genome sequences from eight Middle Eastern populations (Almarri et al., 2021). Phased VCF files of Middle Eastern populations were obtained from Almarri et al, and underwent QC using *plink* including confirming genotyping call rate of greater than 95%, removing variants with a minor allele frequency <0.01 (1%), and removing variants with Hardy-Weinberg equilibrium p-values <1110^-6^. The resulting plink files were merged with HapMap3 genotypes, resulting in a reference panel of 1125 individuals and 10,243 SNPs. Ancestry inference in the UK Biobank population was performed using the software KING (default settings) (Manichaikul et al., 2010), based on a genomic reference panel constructed from HapMap3 and MENA populations. KING first implements multidimensional scaling (MDS) to identify the top 10 ancestry components for each individual. The MDS components were projected onto the HapMap3 + MENA reference panel component space, and genetic ancestry was inferred using support vector machines from all 10 MDS components via the *e1071 (Dimitriadou et al., 2009)* package in R. Additionally, the probability of each individual belonging to each ancestry in the reference panel was extracted. We excluded participants in the British, German, and Indian comparison groups who were considered to have at least an 80% probability of being genetically classified as MENA based on genomic data from (Almarri et al., 2021). We assessed minor allele frequency differences across populations of the lead SNPs of genome-wide significant loci from a recent ADRD GWAS meta-analysis (N = 74,046) (Lambert et al., 2013) We included 18/19 loci/SNPs available within the UKB.

Though larger and more recent AD GWAS studies have been published (Andrews et al., 2023), we used the latest available GWAS that did not include UKB to avoid sample overlap. Given its importance in ADRD risk, we also analyzed ApoE haplotypes.

We also assessed family history for several major chronic disorders. We report the prevalence of combined maternal, paternal, or sibling family history of heart disease, stroke, chronic bronchitis/emphysema, hypertension, diabetes, AD/dementia, Parkinson’s disease, severe depression, and cancers of the lung, bowel, breast, and prostate.

### Statistical analysis

#### Nonimaging

The Shapiro-Wilk test was used to assess normality of the data. Continuous variables that did not follow a normal distribution were compared across groups using the Wilcoxon rank-sum test, while normally distributed continuous variables were compared using the Student’s t-test. For categorical variables, the Chi-squared test was applied unless any group sample size was below 5, in which case Fisher’s exact test was used. For AD risk SNPs, minor allele frequency was calculated in addition to performing Chi-squared tests. For diseases and hearing, eye, and mouth conditions, sex-stratified logistic regressions were performed to account for age, TDI, and age interaction with birth country group to ensure the prevalences were adjusted for. Benjamini-Hochberg corrections for multiple testing were applied separately within each MENA comparison (MENA vs British, MENA vs Germany, and MENA vs India) for environmental variables, family history, and AD SNPs. Additional corrections were performed for self-reported lifestyle, objective measures, blood biochemistry and assays, conditions, and diseases each of which was further stratified by sex.

#### Imaging

For quantitative measures including average thickness, hippocampal volume, white matter hyperintensity volume, global FA, and global MD, linear regression models were run with an age by birth country group interaction and sex and scan site as covariates. Intracranial volume (ICV) was also included as a covariate in the average hippocampal volume and white matter hyperintensity volume models. Logistic regression was used to assess the presence of total microbleeds (definite+probable+possible), where the classification of having CMBs was determined by the presence of at least one CMB. These models included an interaction between birth country group and age as well as sex as a covariate. Benjamini & Hochberg correction for multiple testing was performed across all imaging measures for each covariate. Given (1) the similarity we found across the nonimaging features between MENA and India as well as between the British and German groups and (2) the substantially smaller N available across imaging features, we opted to combine the similar groups across the imaging comparisons. The original grouping results are still available in the supplementary materials (**Tables S27-S29**). Post hoc analysis on regional MD measures was performed to investigate whether there was a localized effect.

Supplementary materials can be found at: https://brainescience.shinyapps.io/adrd_mena_ukb/.

## Results

### Population characteristics

Overall, 49% of the study sample were female and the mean age was 54.28±8.37. A total of 3,552 participants were classified as MENA (38%F; mean age 52.54 ± 7.99), 1,925 as British (56%F; mean age 54.60 ± 7.69), 1,097 as German (75%F; mean age 55.78 ± 9.46), and 2,935 as Indian (47%F; mean age 55.63 ± 8.43) (**Figure 1A**; **Figure 2A**, **Table 1**). The MENA group included participants from 34 different birth countries. The five most represented countries were Pakistan (n=1,349), Iran (n=505), Iraq (n=312), Egypt (n=155), Turkey (n=145) and Cyprus (n=145). The remaining countries had between 1 and 113 participants. The British group comprised individuals from 24 birth countries, with the top five being Germany (n=839), India (n=378), Malta (n=194), Cyprus (n=131), and Egypt (n=94). All other countries contributed between 1 and 47 participants. These distributions are illustrated in **Figure 1B** and detailed further in **Table S1**.

**Figure 2.**
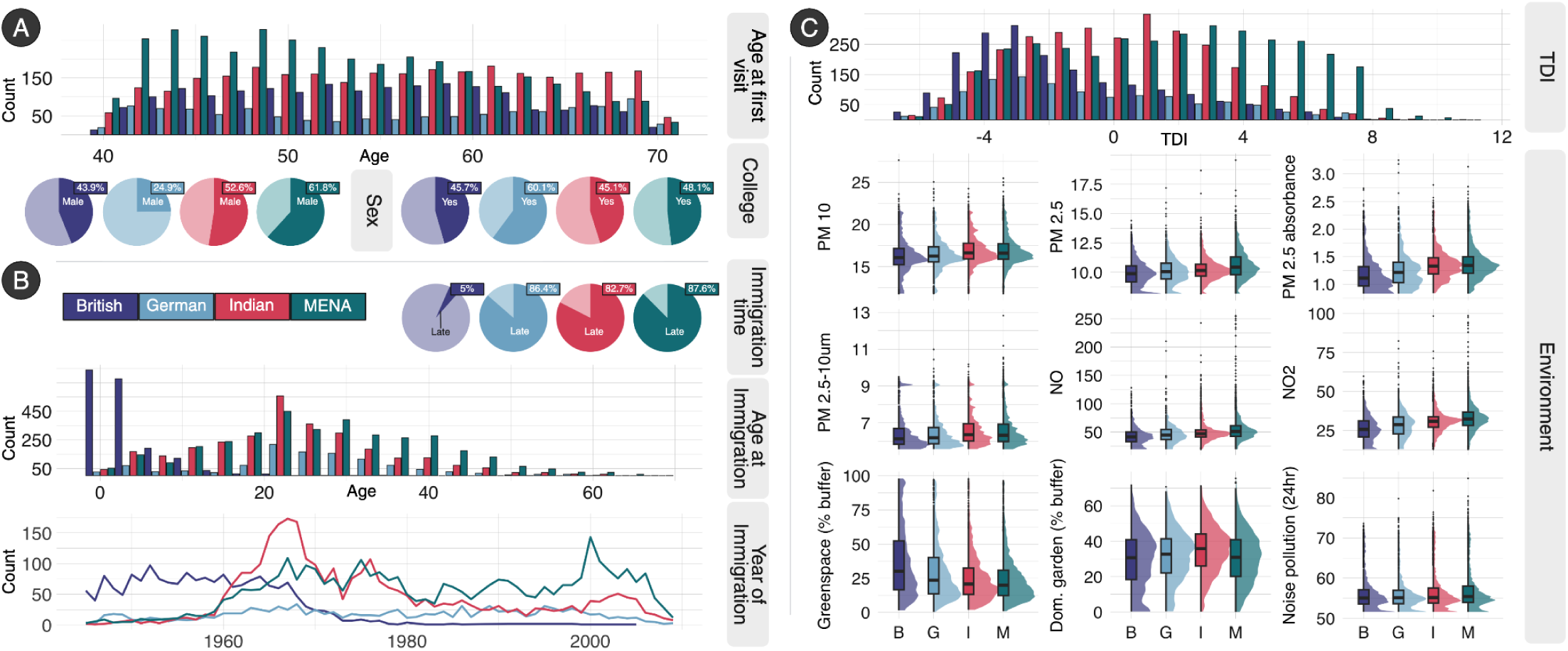
Population Demographics. **A.** Distribution of age at baseline visit, sex, and college education. **B.** Age and year of immigration as well as distributions of early (before the age of 12) migrants and late migrants (after the age of 12). **C.** Area deprivation (as measured by TDI) and environmental factors across populations. The MENA population were generally younger, had a higher proportion of males, and lived in areas with more deprivation and pollution.

**Table 1.** Demographic characteristics.

| Characteristic | British<br>N = 1925<sup>†</sup> | Germany<br>N = 1097<sup>†</sup> | India<br>N = 2935<sup>†</sup> | MENA<br>N = 3552<sup>†</sup> | Overall<br>N = 9509<sup>†</sup> |
| --- | --- | --- | --- | --- | --- |
| Sex |  |  |  |  |  |
| Female | 1080 (56%) | 824 (75%) | 1392 (47%) | 1357 (38%) | 4653 (49%) |
| Male | 845 (44%) | 273 (25%) | 1543 (53%) | 2195 (62%) | 4856 (51%) |
| Age at first visit | 54.60 (7.69) | 55.78 (9.46) | 55.63 (8.43) | 52.54 (7.99) | 54.28 (8.37) |
| Age immigrated | 3.56 (4.66) | 24.98 (11.38) | 22.81 (11.46) | 26.79 (12.90) | 20.83 (14.05) |
| Townsend deprivation index | -1.38 (3.00) | -0.62 (3.26) | -0.03 (2.93) | 1.46 (3.61) | 0.18 (3.43) |
| Attended college | 878 (46%) | 657 (60%) | 1276 (45%) | 1645 (48%) | 4456 (48%) |
| ApoE4 counts |  |  |  |  |  |
| 0 | 1360 (71%) | 814 (74%) | 2410 (82%) | 2957 (83%) | 7541 (79%) |
| 1 | 513 (27%) | 264 (24%) | 495 (17%) | 564 (16%) | 1836 (19%) |
| 2 | 52 (2.7%) | 19 (1.7%) | 30 (1.0%) | 31 (0.9%) | 132 (1.4%) |
| Hypertension | 550 (29%) | 329 (30%) | 1361 (46%) | 1414 (40%) | 3654 (38%) |
| Diabetes | 139 (7.2%) | 63 (5.7%) | 767 (26%) | 833 (23%) | 1802 (19%) |
| Hypercholesterolemia | 266 (14%) | 176 (16%) | 1047 (36%) | 1102 (31%) | 2591 (27%) |
| On hypertension medication | 258 (13%) | 161 (15%) | 862 (29%) | 723 (20%) | 2004 (21%) |
| On cholesterol medication | 223 (12%) | 118 (11%) | 819 (28%) | 815 (23%) | 1975 (21%) |
| Systolic blood pressure | 133.92 (17.24) | 134.45 (19.52) | 136.72 (18.80) | 132.56 (17.41) | 134.34 (18.15) |
| Diastolic blood pressure | 81.56 (9.89) | 80.91 (10.28) | 82.65 (10.04) | 81.57 (10.01) | 81.83 (10.04) |
| Body mass index | 27.06 (4.87) | 25.63 (4.45) | 26.83 (4.03) | 28.22 (4.61) | 27.26 (4.56) |
| Smoking |  |  |  |  |  |
| Current | 227 (12%) | 151 (14%) | 196 (6.7%) | 698 (20%) | 1272 (13%) |
| Previous | 710 (37%) | 461 (42%) | 300 (10%) | 766 (22%) | 2237 (24%) |
| Never | 985 (51%) | 481 (44%) | 2410 (83%) | 2061 (58%) | 5937 (63%) |
| Alcohol |  |  |  |  |  |
| Frequent | 496 (26%) | 250 (23%) | 268 (9.2%) | 181 (5.1%) | 1195 (13%) |
| Occasional | 899 (47%) | 523 (48%) | 723 (25%) | 576 (16%) | 2721 (29%) |
| Infrequent | 529 (27%) | 323 (29%) | 1930 (66%) | 2773 (79%) | 5555 (59%) |
| Diet quality score | 49.58 (13.45) | 53.64 (13.03) | 51.94 (11.85) | 47.59 (13.67) | 50.03 (13.20) |
| Physical activity |  |  |  |  |  |
| High | 668 (42%) | 400 (43%) | 752 (34%) | 836 (31%) | 2656 (36%) |
| Moderate | 635 (40%) | 420 (46%) | 952 (43%) | 1078 (40%) | 3085 (41%) |
| Low | 284 (18%) | 103 (11%) | 513 (23%) | 793 (29%) | 1693 (23%) |
| Sleep score |  |  |  |  |  |
| Healthy | 691 (36%) | 406 (37%) | 955 (33%) | 967 (27%) | 3019 (32%) |
| Intermediate | 1113 (58%) | 628 (57%) | 1727 (59%) | 2169 (61%) | 5637 (59%) |
| Poor | 121 (6.3%) | 63 (5.7%) | 253 (8.6%) | 416 (12%) | 853 (9.0%) |
| Self rated health |  |  |  |  |  |
| Excellent | 406 (21%) | 201 (18%) | 253 (8.6%) | 318 (9.0%) | 1178 (12%) |
| Good | 1071 (56%) | 664 (61%) | 1480 (50%) | 1573 (44%) | 4788 (50%) |
| Fair | 369 (19%) | 194 (18%) | 913 (31%) | 1116 (31%) | 2592 (27%) |
| Poor | 74 (3.8%) | 30 (2.7%) | 235 (8.0%) | 460 (13%) | 799 (8.4%) |
| Do not know | 5 (0.3%) | 8 (0.7%) | 43 (1.5%) | 62 (1.7%) | 118 (1.2%) |
| Prefer not to answer | 0 (0%) | 0 (0%) | 10 (0.3%) | 22 (0.6%) | 32 (0.3%) |

n (%); Mean (SD)

The rate of college attendance in MENA (48.1%) was significantly less than those from Germany (60.1%; χ^2^=47.6; *p*=5.3×10^-12^), significantly greater than those from India (45.1%; χ^2^=5.7; *p*=1.7×10^-2^), and did not differ significantly from the British group (45.7%; *p*=8.3×10^-2^) (**Figure 2A**). The mean age of immigration was significantly higher in MENA (26.79±12.90) compared to all other British (3.56±4.66; *p*<0.001), German (24.98±11.38; *p*=1.2×10^-3^), and Indian (22.81±11.46; *p*=2.9×10^-36^) populations. The British group was predominantly composed of early-life migrants, with 95% having migrated before the age of 12. In contrast, 80% of participants in the MENA, German, and Indian groups migrated after age 12 (**Figure 2B**). Material deprivation, as measured by the Townsend Deprivation Index, was highest in the MENA group (mean score: 1.5 ± 3.6), indicating above-average levels of deprivation. This was significantly higher than all other groups including the Indian group which had a mean score near zero (0 ± 2.9; *z*=17.0; *p*=1.2×10^-64^), suggesting average deprivation levels relative to the general population and also the British (-1.4 ± 3.0; *z=*27.5; *p*=3.2×10^-166^) and German (-0.6 ± 3.3; *z*=-16.6; *p*=4.9×10^-62^) groups which had negative scores, reflecting relatively more affluent socioeconomic conditions. Levels of environmental pollution, including coarse particulate matter (PM₁₀), fine particulate matter (PM₂.₅), nitrogen oxide (NO), and nitrogen dioxide (NO₂), were generally higher in MENA compared to the Indian, German, and British groups. The British group resided in areas with the lowest overall pollution levels (**Figure 2C**). The percentage of greenspace and domestic garden buffers around household location was also generally lower in MENA compared to all other populations. Similarly, 24 hour noise pollution was highest in MENA compared to all other groups (**Table S13**).

### Family History and AD Genetic Risk Factors

Rates of a family history of heart disease and stroke were similar across all four populations. Family history of high blood pressure in MENA (54%) was similar to Indian (55.4%) populations but significantly higher than German (49.5%; χ^2^=6.5; *q*=1.8×10^-2^) and British (48.8%; χ^2^=13.3; *q*=3.6×10^-4^) populations. Family history of diabetes was significantly lower in MENA (43.3%) compared to the Indian (48.1%; χ^2^=14.4; *q*=4.4×10^-4^) group but was nearly double the rate compared to German (22.5%; χ^2^=152.6; *q*=5.7×10^-34^) and British (20.8%; χ^2^=275.2; *q*=9.9×10^-61^) populations. Family history of AD/dementia in MENA (6.4%) was nearly half that of German (11.9%; χ^2^=34.5; *q*=1.7×10^-8^) and British (11.9%; χ^2^=49.7; *q*=7.2×10^-12^) populations but did not differ significantly from the Indian group (5.5%) Generally, family history of cancer was lower in MENA compared to British and German populations but higher compared to Indian populations (**Figure 3A; Table S14**). The MENA population had mothers whose age at death was significantly lower (68.35±14.55) compared to Indian (69.94±14.46; *z*=-3.5; *p*=4×10^-4^), British (73.94±13.79; *z*=-9.7; *p*=3.2×10^-22^), and German (74.04±14.64; *z*=8.8; *p*=1.1×10^-18^) populations (**Figure 3B; Table S12**).

**Figure 3.**
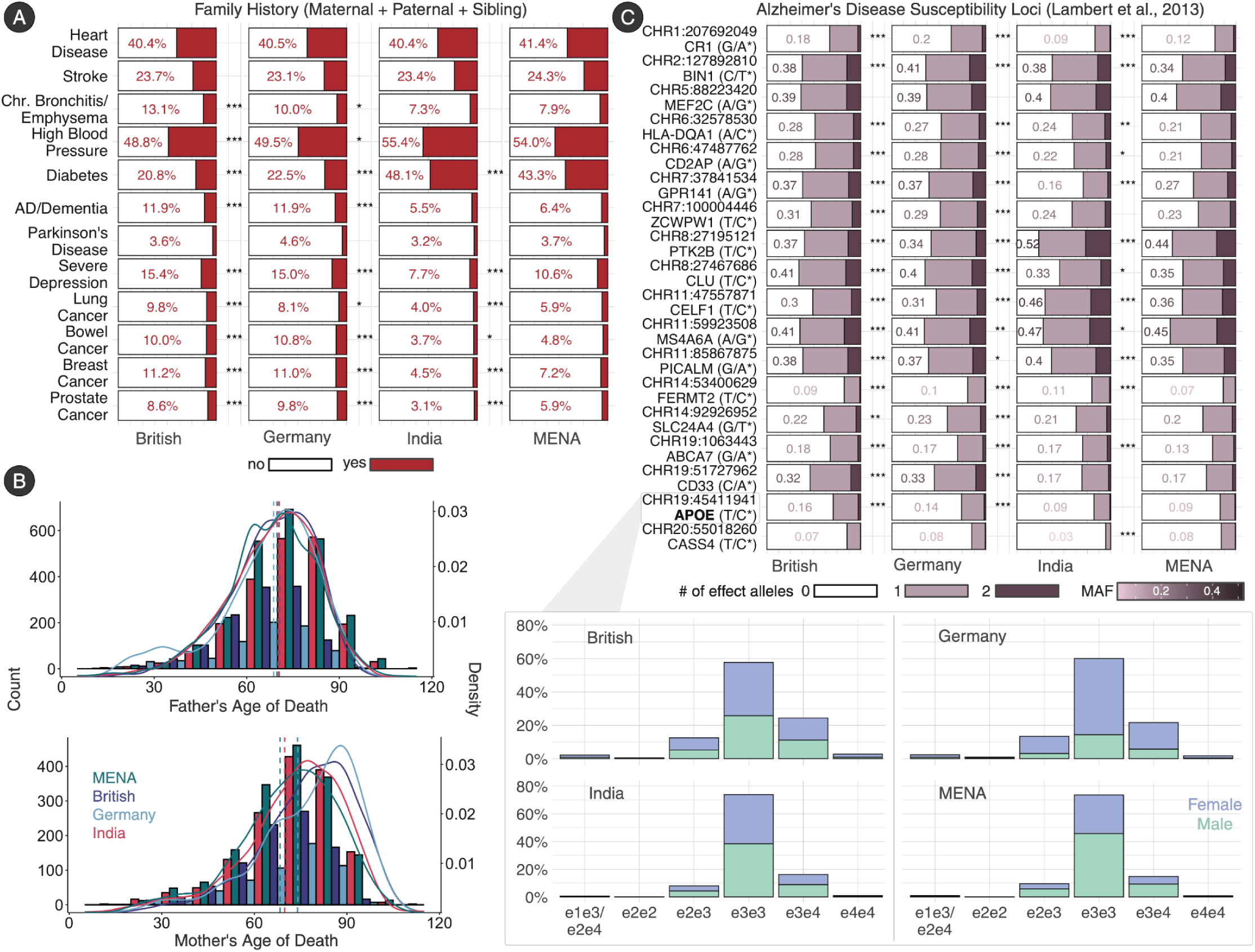
Genetics. **A.** Combined maternal, paternal, and sibling family history comparisons between MENA and British, German, and Indian populations. **B.** Maternal and paternal age of death distributions across the four populations. **C.** MENA comparisons of AD susceptibility loci frequencies from Lambert et al., 2013. ApoE genotype distributions are also shown across sex for the four populations.

The number of effect alleles from susceptibility loci implicated in Alzheimer’s disease differed most when comparing MENA to British and German populations with significantly lower frequency in 13/18 susceptibility loci tested, and significantly higher frequency in 3/18 AD loci. Half (9/18) of the loci occurred in higher frequency in Indians compared to MENA and 4/18 had significantly lower frequency in Indians compared to MENA. Notably, MENA had significantly lower frequency of the ApoE isoform e4 (minor allele frequency (MAF):0.09) compared to Germany (MAF:0.14; χ^2^=45.8; *q*=4×10^-11^), and British (MAF:0.16; χ^2^=129.2; *q*=5.6×10^-29^) populations (**Figure 3C; Table S15**). The frequency of the e4 isoform did not differ in MENA compared to Indian populations.

### Lifestyle factors

#### Diet

Overall, the MENA group had significantly lower diet quality scores among both females and males compared to all 3 other populations. Among females, the mean diet quality score (max 100) was 49.96 ± 13.14, compared to 53.18±11.06 in the Indian group (*z*=-7.2; *q*=5.3×10^-12^), 54.08 ± 12.75 in the German group (*z*=-7.2; *q*=2.1×10^-12^), and 50.78±12.56 in the British group (*z*=-2.2; *q*=3.5×10^-2^). Components that contributed to lower scores in MENA females compared to British females were in categories that represented favorable intake of whole grains, fish, dairy, vegetable oil, and refined grains, however MENA females did have significantly higher scores in categories that represented favorable intake of fruits, vegetables, processed meat, unprocessed meat, and sugary foods/drinks. Components that contributed to lower total scores in MENA females compared to German females were in categories that represented favorable intake of whole grains, fish, dairy, and refined grains, while the only category that MENA females had better scores in was favorable intake of processed meats. Favorable intake of vegetable oil, processed and unprocessed meats contributed to a higher overall diet score in Indian females compared to MENA females but MENA females did have higher favorable intake of whole grains, fish, and sugary foods/drinks compared to the Indian group. Similarly, among males, the MENA group had a score of 46.12±13.78, which was lower than scores in the Indian (50.82±12.41; *z*=-10.8; *q*=1.6×10^-26^), German (52.29±13.76; *z*=6.8; *q*=6.3×10^-11^), and British (48.05±14.37; *z*=-3.7; *q*=3.7×10^-4^) groups. Components that contributed to lower total diet scores in MENA males compared to British males were in categories that represented favorable intake of whole grains, fish, dairy, vegetable oil, and refined grains, however MENA males did have more favorable scores in the fruit, vegetable, processed meat, and unprocessed meat categories. Components that contributed to lower total diet scores in MENA males compared to German males included whole grains, fish, dairy, vegetable oil, and refined grains, while the only category where MENA males had more favorable scores was in processed meat intake. Higher total diet scores in Indian males compared to MENA males were represented by more favorable intake in vegetables, whole grains, vegetable oil, refined grains, processed and unprocessed meats, however MENA males scored higher in fruit, fish, and dairy intake. In both males and females, rates of fish oil consumption was significantly lower in MENA compared to both the British and German populations but did not differ significantly compared to the Indian group. (**Figure 4A; Tables S4-S5; S16-S17**).

**Figure 4.**
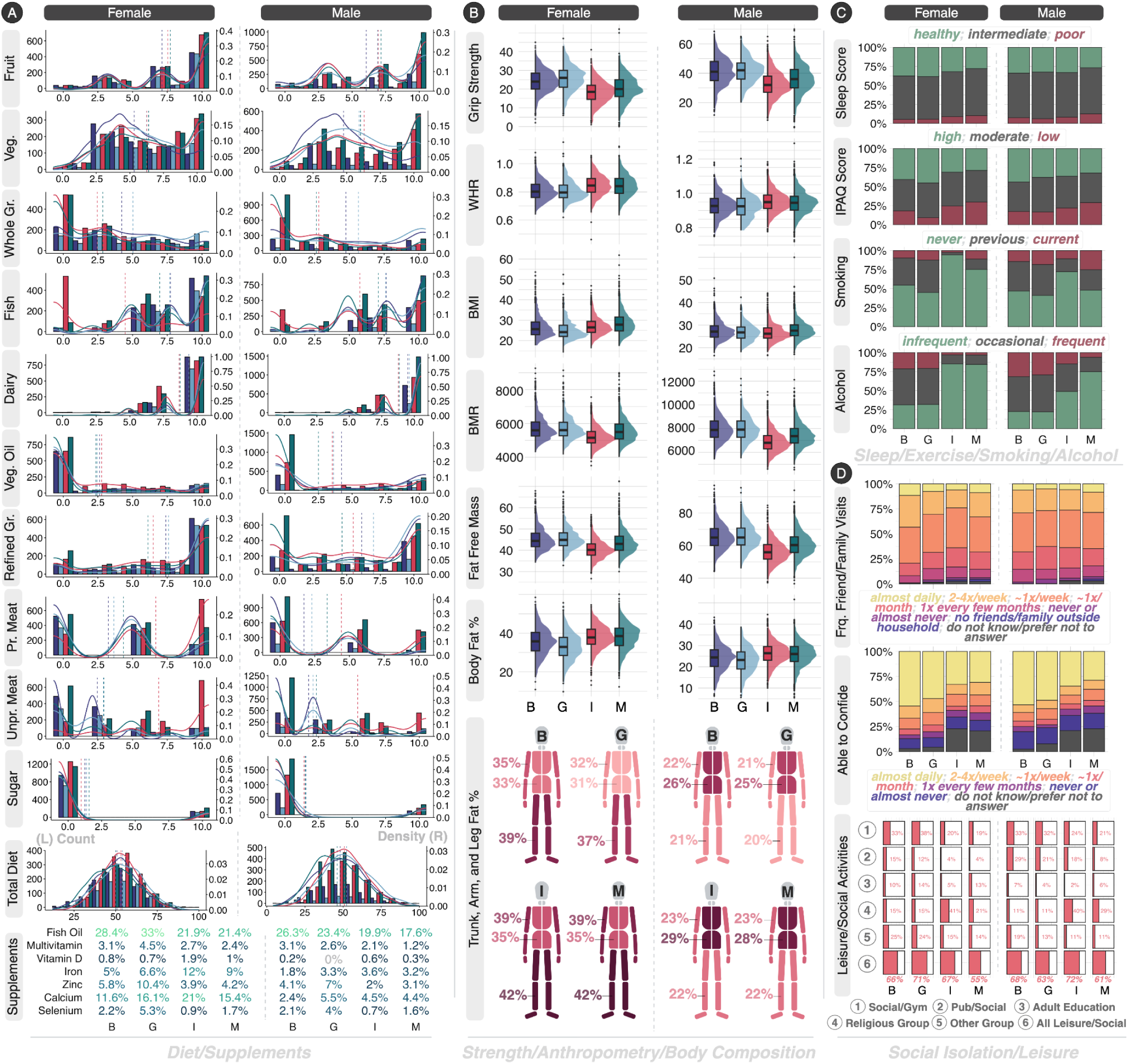
Lifestyle factors. **A.** Distribution across the four populations of 10 dietary components (fruit, vegetables, whole grains, fish, dairy, vegetable oil, refined grains, processed meat, unprocessed meat, and sugary food/drink intake) that make up the total diet score across females (L) and males (R). Y-axis on the left displays the raw counts and the y-axis on the right represents the density. Supplement use prevalence across the four populations in females (L) and males (R). **B.** Strength, anthropometry, and body composition distributions across the four populations in females (L) and males (R). **C.** Distributions of sleep, exercise, smoking, and alcohol use scores and variables across the four populations in females (L) and males (R). **D.** Answers to social isolation questionnaires including frequency of friend/family visits and how often the participant is able to confide in someone in females (L) and males (R). Distributions of leisure activities are also included. *B: British; G: German; I: Indian; M: MENA; Veg: vegetables; Gr: grains; Pr: processed; Unpr: unprocessed; WHR: waist-to-hip ratio; BMI: body mass index; IPAQ: International Physical Activity Questionnaire; Frq: frequency*

#### Body composition

Both MENA females and males had significantly lower grip strength compared to British and German populations. Measured in units of force production, MENA females had a mean grip strength of 20.42±6.77kg compared to 25.71±6.72kg in German females (*z*=-16.7; *q*=1.0×10^-61^), and 24.38±5.92kg in British females (*z*=-14.5; *q*=1.6×10^-46^). However, MENA females had significantly higher grip strength compared to Indian females, who had the weakest grip strength at 18.42±5.89 kg (*z*=8.2; *q*=1.0×10^-15^). There was a similar pattern among males: MENA males had lower grip strength (35.79±9.56kg) compared to German (41.77±8.34kg; *z*=9.62; *q*=7.4×10^-21^) and British (41.14±9.24kg; *z*=-13.40; *q*=2.8×10^-40^) males but higher grip strength compared to Indian males (32.63±8.59kg; *z*=10.80; *q*=1.2×10^-26^), who had the weakest grip strength. In terms of body composition, both MENA females and males had worse outcomes compared to British and German populations. MENA females for example had a waist to hip ratio (WHR) of 0.84±0.08 and total body fat percentage (BF) of 38% which was significantly higher than both German (WHR: 0.81±0.07; *z*=11.4; *q*=5.1×10^-30^; BF: 33%; *z*=15.9; *q*=1.5×10^-56^) and British (WHR: 0.81±0.07; *z*=11.4; *q*=1.7×10^-29^; BF: 36%; *z*=8.8; *q*=3.4×10^-18^) females. The higher body fat in MENA compared to British and German females seemed to be most apparent in arm fat (4-7% mean difference), followed by leg fat (3-5% mean difference), and finally trunk fat (2-4% mean difference). While MENA females did not differ significantly compared to Indian females in WHR or BF, MENA females did have higher BMI (28.62±5.43) but also higher whole body fat free mass (FFM) (43.62±5.27kg) compared to Indian females (BMI: 27.08±4.42; *z*=7.4; *q*=3×10^-13^; FFM: 40.56±4.27; *z*=15.6; *q*=1.1×10^-53^). Similar to females, MENA males had higher WHR (0.95 ± 0.06) and BF (26%) compared to German (WHR: 0.92±0.07; *z*=-4.91; *q*=1.1×10^-6^; BF: 23%; *z*=-7.47; *q*=2.3×10^-13^) and British (WHR: 0.93±0.06; *z*=6.38; *q*=2.7×10^-10^; BF: 24%; *z*=7.5; *q*=1.4×10^-13^) males. Differences in body fat differed most in the trunk (2-3% mean difference), followed by leg and arm fat (1-2% mean difference) when comparing MENA to British and German populations. MENA males had lower WHR (0.95±0.06), higher whole body FFM (60.82±7.28 kg), but higher BMI (27.96±4.01) compared to Indian males (WHR: 0.95±0.06; *z*=-2.81; *q*=7.9×10^-3^; FFM: 56.35±6.81; *z*=18.2; *q*=1.3×10^-72^; BMI: 26.63±3.64; *z*=10.70; *q*=2×10^-26^). MENA populations had lower basal metabolic rate (BMR) compared to British and German populations but higher BMR compared to Indian populations. In females, BMR for MENA was 5583±696 kJ/day which was significantly greater than Indian females (5210±549kJ/day; *z*=14.5; *q*=1.1×10^-46^), but less than German (5702±649 kJ/day; *z*=-4.6; *q*=5.2×10^-6^) and British (5731±707 kJ/day; *z*=5.1; *q*=3.3×10^-7^) females. Similarly in males, BMR for MENA was 7443±949 kJ/day which was significantly greater than Indian males (6884±869 kJ/day; z=17.8; q=2.4×10-70), but less than German (8001±1061 kJ/day; z=8.32; q=3.2×10-16) and British (8002±1083 kJ/day; z=-13; q=6.5×10-38) males. (**Figure 4B; Tables S6-S7; S18-S19**).

#### Sleep, physical activity, smoking, & alcohol

The MENA population had worse sleep scores (based on a composite self report measure) compared to the other populations in both females and males, having the highest proportion of “poor” sleep (females: MENA: 10%; British: 5.3%; German: 5.6%; Indian: 9%; males: MENA: 13%; British: 7.6%; German: 6.2%; Indian:8.3%) and the lowest proportion of “healthy” sleep (females: M:28%; B:38%; G:38%; I:32%; males: M:27%; B:34%; G:33%; I:33%). Sleep categorizations differed significantly in MENA females compared to British (χ^2^=37.3; *q*=1.8×10^-8^) and German (χ^2^=33.7; *q*=1.2×10^-7^) females. In MENA males, sleep categorizations differed significantly compared to British (χ^2^=25; *q*=6.9×10^-6^), German (χ^2^=11.4; *q*=5.8×10^-3^), and Indian (χ^2^=28.6; *q*=1.7×10^-6^) males. Similarly the MENA population had worse physical activity scores compared to the other populations in both females and males, having the highest proportion of “low” physical activity scores (females: MENA: 30%; British: 18%; German: 9.3%; Indian: 25%; males: MENA: 29%; British: 17%; German: 17%; Indian: 22%) and the lowest proportion of “high” physical activity scores (females: MENA: 29%; British: 41%; German: 45%; Indian: 31%; males: MENA: 32%; British: 44%; German: 38%; Indian: 36%). Physical activity scores differed significantly comparing MENA females to British (χ^2^=43.6; *q*=9.5×10^-10^) and German (χ^2^=112.4; *q*=2.4×10^-24^) females whereas statistical differences in males were found comparing MENA to British (χ^2^=46.6; *q*=1.8×10^-10^), German (χ^2^=15.6; *q*=1×10^-3^), and Indian (χ^2^=18.6; *q*=1.9×10^-4^) populations. Smoking status significantly differed between MENA and all other populations. Overall, MENA females had higher prevalence of current (11%) and previous (14%) smoking and lower prevalence of never having smoked (75%) compared to Indian females (current:2.3%, previous:3.2%; never:95%; χ^2^=198.2; q=6.9×10^-43^). Similar rates were found between MENA females and British (9.8%) and German (12%) females for current smoking but British and German females had a higher prevalence of previously smoking (British: 36%; German: 43%) and a lower prevalence of never having smoked (British: 55%; German: 45%) compared to MENA (British: χ^2^=162.8; *q*=4.4×10^-35^; German: χ^2^=248.9; *q*=8.8×10^-54^). MENA males had the highest rates of currently smoking (25%) compared to British (14%), German (18%), and Indian (11%) males. Similar to females, MENA males had higher rates of previously smoking (27%) and lower rates of never having smoked (48%) compared to Indian males (previous:17%; never:72%; χ^2^=227.3; q=4.5×10^-49^). MENA males had slightly higher rates of never having smoked (48%) compared to British (47%) and German (41%) males and lower rates of previously having had smoked (27%) compared to British (39%) and German (41%) males (British: χ^2^=61.4; *q*=1.3×10^-13^; German: χ^2^=23.8; *q*=2.3×10^-5^). Alcohol intake among MENA females was significantly lower (frequent:3.3%; occasional:12%; infrequent:85%) compared to British (frequent: 21%; occasional: 47%; infrequent: 31%; χ^2^=716.4; *q*=8.3×10^-155^) and German (frequent: 21%; occasional: 47%; infrequent: 32%; χ^2^=623.9; q=1×10^-134^) females but did not differ compared to Indian females (frequent: 2.8%; occasional: 12%; infrequent: 85%; *ns*). MENA males had the lowest prevalences of alcohol intake (frequent: 6.3%; occasional: 19%; infrequent: 75%) compared to British (frequent: 32%; occasional: 46%; infrequent: 22%; χ^2^=737.9; *q*=1.7×10^-159^), German (frequent: 29%; occasional: 49%; infrequent: 22%; χ^2^=341; *q*=2.6×10^-73^), and Indian (frequent: 15%; occasional: 36%; infrequent: 49%; χ^2^=266.7; *q*=1.9×10^-57^) males. (**Figure 4C; Tables S4-S5; S16-S17**).

#### Social isolation & loneliness

A higher proportion of MENA females (12%) and males (14%) were considered “socially isolated” (based on a summary score taking into account living alone, infrequent family/friend visits, and whether or not one engages in leisure/social activities) compared to Indian females (9.4%; χ^2^=5.3; *q*=4.6×10^-2^) and males (8%; χ^2^=30.9; *q*=8.9×10^-8^). Female and male people from MENA were also generally considered more lonely compared to other populations. In females, 9% of MENA considered themselves lonely which was significantly higher than British (5.2%; χ^2^=11.3; *q*=1.2×10^-3^) and German (5.4%; χ^2^=8.2; *q*=6.3×10^-3^) but not Indian (9.7%; *ns*) females. In males, 10% of MENA considered themselves lonely, significantly higher than British (7.6%; χ^2^=5.1; *q*=3.2×10^-2^), German (4.8%; χ^2^=7.7; *q*=9.2×10^-3^), and Indian (7.9%; χ^2^=5.1; *q*=3.5×10^-2^) males. Importantly, MENA and Indian populations had higher proportions of non-substantive responses when answering questions that went into calculating the loneliness score across males and females. For example, for the question “how often are you able to confide in someone close to you” 21.6% of MENA and 22% of Indian populations responded with “do not know” or “prefer not to answer” compared to 2.7% of British and 5.2% of German populations. Similarly for “do you often feel lonely?”, 6.5% of MENA and 6% of Indians responded “do not know” or “prefer not to answer” compared to only 0.9% of British and 1.7% of German participants. Overall, MENA had the lowest self-rated health among both males and females compared to all other populations, though to a lesser degree compared to Indian populations. In females, 50.1% of MENA rated their health as “excellent” or “good” compared to 79% of British, 80% of German, and 56% of Indian females. MENA females instead had the largest proportion of “fair” or “poor” ratings (48%) compared to 20.7% of British, 19.8% of German, and 41.4% of Indian females. Similarly in males, 55.5% of MENA rated their health as “excellent” or “good” compared to 74% of British, 77% of German, and 62% of Indian males. MENA males had the largest proportion of “fair” or “poor” ratings (42%) compared to 26% of British, 23.6% of German, and 36.7% of Indian males. (**Figure 4D; Tables S4-S5; S16-S17**).

### Disease Prevalence

Overall, MENA had higher rates of cardiovascular disease compared to British and German populations. Both females and males had more than double the odds of coronary artery disease compared to British (F: OR=2.73; *q*=2.8×10^-7^; M: OR=2.73; *q*=9.5×10^-11^) and German (F: OR=2.59; *q*=8.4×10^-6^; M: OR=2.02; *q*=9.3×10^-3^) populations. MENA females also had nearly 4 times the odds of having heart failure compared to German females (OR=3.95; *q*=3.2×10^-2^). MENA females had over 3 times the odds and MENA males had over 2 times the odds of having high cholesterol compared to British females (F: OR=3.51; *q*=5.5×10^-19^; M: OR=2.71; *q*=1.5×10^-15^). MENA females had higher odds of hypertension compared to British (OR=1.76; *q*=3.5×10^-8^) and German (OR=2.56; *q*=2.6×10^-14^) females. While German females had statistically higher systolic blood pressure at baseline assessment (*z*=-2.5; *q*=1.3×10^-2^) compared to MENA females, a higher proportion of MENA females (17%) were on blood pressure medication compared to German females (11%). MENA males had higher odds of hypertension compared to the British males (OR=1.62; *q*=1.6×10^-6^). Similarly, while British males had statistically higher systolic (*z*=-5.25; *q*=1.8×10^-7^) and diastolic (*z*=-2.56; *q*=1.1×10^-2^) blood pressure as baseline assessment, a higher proportion of MENA males (23%) were on blood pressure medication compared to British males (15%). Similarly, MENA had higher rates of metabolic dysfunction. Strikingly, MENA females were 4-6 times more likely to have diabetes compared to British (OR=4.69; *q*=2.1×10^-20^) and German (OR=5.96; *q*=1.8×10^-17^) populations. MENA males were 2-4 times more likely to have diabetes compared to British (OR=2.58; *q*=1.4×10^-11^) and German (OR=4.05; *q*=5.8×10^-6^) populations. MENA females were 1-3 times more likely to be obese compared to British (OR=1.69; *q*=3.4×10^-7^), German (OR=3.11; *q*=5×10^-19^), and Indian (OR=1.79; *q*=1.5×10^-9^) females. MENA males were 1-2 times more likely to be obese compared to German (OR=1.51; *q*=4.2×10^-2^) and Indian (OR=2.08; *q*=1.9×10^-14^) populations.

MENA females had higher odds of depression compared to German (OR=1.59; *q*=4.1×10^-2^) and Indian (OR=1.53; *q*=4.3×10^-2^) females. MENA females also had twice the odds of having anxiety compared to Indian females (OR=2.12; *q*=7.3×10^-4^). MENA males had twice the odds of depression compared to Indian males (OR=2.20; *q*=9.1×10^-4^) but had 38% lower odds compared to British males (OR=0.62; *q*=2.6×10^-2^). MENA females had higher rates of headaches or migraines compared to all three British (OR=1.64; *q*=5.7×10^-6^), German (OR=2.26; *q*=5.4×10^-11^), and Indian (OR=1.42; *q*=7.3×10^-4^) populations. The rates for headaches or migraines did not statistically differ between MENA males and British and German males, but were higher compared to Indian (OR=1.53; *q*=1.1×10^-3^) males. MENA females had 3 times the odds of having sleep apnea compared to German females (OR=3.14 *q*=4.8×10^-2^) but there weren’t significant differences between the other populations or among males. Interestingly, MENA had consistently lower rates of cancer compared to the British and German populations, but no differences were reported compared to Indians. In females, odds of cancer were 49% (*q*=7.2×10^-4^) lower in MENA compared to British and 37% (*q*=4.1×10^-2^) lower in MENA compared to Germans. In males, odds of cancer were 63% (*q*=1.9×10^-4^) lower in MENA compared to British and 64% (*q*=9.9×10^-3^) lower in MENA compared to Germans.

MENA males had lower odds of hearing difficulties than British (OR=0.57; *q*=9.2×10^-7^) and German (OR=0.61; *q*=1×10^-2^) males. MENA females had over 8 times the odds of being diagnosed with a diabetes related eye condition compared to British females (OR=8.24; *q*=5.1×10^-7^) and over 2 times the odds compared to German females (OR=2.40; *q*=2.1×10^-3^). MENA males had nearly 6 times the odds of diabetes related eye conditions compared to British males (OR=5.91; *q*=3.9×10^-6^). MENA females also had higher odds of cataracts compared to British (OR=3.28; *q*=2.6×10^-3^) and German (OR=2.54; *q*=3×10^-2^) females. MENA also had higher odds of indicators of periodontal disease such as painful and bleeding gums compared to British and German populations. MENA females had higher odds of painful gums compared to British females (OR=2.09; *q*=2.8×10^-4^) and higher odds of painful (OR=1.92; *q*=2.9×10^-3^) and bleeding (OR=1.48; *q*=3×10^-3^) gums compared to German females. Similarly, MENA males had higher odds of both painful and bleeding gums compared to British (painful: OR=1.96; *q*=6.4×10^-3^; bleeding: OR=1.4; *q*=1.2×10^-2^) and German (painful: OR=2.62; *q*=4.8×10^-2^; bleeding: OR=1.92; *q*=9.7×10^-3^) males (**Figure 5; Tables S22-S23**).

**Figure 5.**
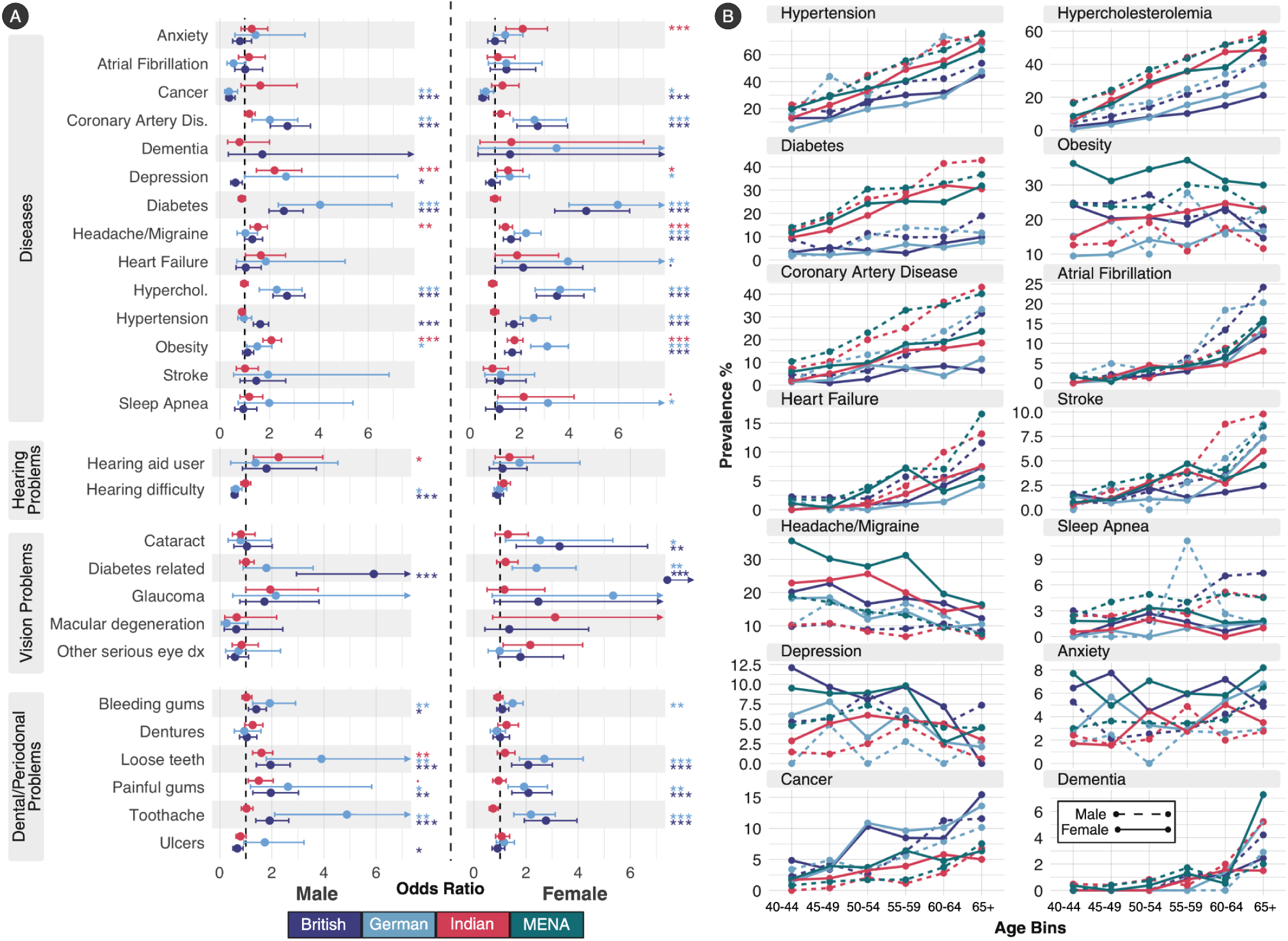
Disease prevalence. **A.** Disease and condition odds ratios comparing MENA to British, German, and Indian populations in males (L) and females (R). **B.** Prevalence of disease binned across 5-year age bins (40-44, 45-49, 50-54, 55-59, 60-64, and 65+ years of age). Solid lines represent females and dashed lines represent males.

### Imaging Features

Global MD was significantly lower in MENA and India combined compared to the European populations (British + Germany; 𝛽 = 0.20; *q* = 0.048). This global effect accounted for the observed regional MD differences, such that no individual regions remained significant after multiple-comparisons correction. Average hippocampal volume was found to be statistically lower in MENA and India combined to the European populations (British + Germany; 𝛽 = 0.19; *q* = 0.044) (**Table S26**).

## Discussion

This study provides the first large-scale characterization of biopsychosocial and neurobiological risk factors for ADRDs among immigrants from MENA countries using data from the UK Biobank. Despite a lower frequency of AD risk SNPs, including ApoE4, MENA immigrants showed a distinctive risk profile marked by greater socioeconomic deprivation, higher exposure to environmental pollutants, adverse lifestyle behaviors, and substantially elevated metabolic and vascular disease burden compared with European immigrant groups. These findings underscore the importance of examining ethnocultural determinants of brain aging and caution against extrapolating dementia risk models derived predominantly from European ancestry populations. This need for context-specific frameworks aligns well with recent regional syntheses, which highlight that dementia risk in the MENA region arises from a convergence of metabolic, vascular, environmental, and healthcare access factors that Western-derived models often fail to capture (Mowafi et al., 2025). Together, these observations emphasize that dementia vulnerability in MENA is shaped by structural and systemic exposures across the life course rather than genetic risk alone (Dhillon & Fazal, 2025a). While our study characterizes the risk architecture of this population rather than dementia incidence itself, these findings provide critical empirical weight to the 2024 Lancet Commission’s assertion that the impact of modifiable risk factors is not uniform across all groups and may have origins at the societal level (Livingston et al., 2024).

Our genetic analyses revealed substantially lower minor allele frequencies of established AD risk variants in MENA participants relative to European comparison groups, in line with previous research (Bowirrat et al., 2000; Sherva et al., 2011). This includes ApoE4, the biggest genetic risk factor in late onset AD (Troutwine et al., 2022), where the minor allele frequency in MENA and India was 9% compared to German and British populations at 14% and 16%, respectively. These finding suggests that the elevated burden of ADRD observed in MENA populations may be driven less by canonical European-derived genetic risk factors and instead may reflect a greater contribution from alternative genetic architectures, non-genetic exposures, or gene-environment interactions that are not captured by current risk models. This discordance between relatively lower genetic risk and substantially higher phenotypic burden also highlights important limitations in extrapolating AD genetics from European populations to individuals of MENA ancestry, as with other non-European ancestries (Farrer et al., 1997; Griswold et al., 2021; Tang et al., 1998). Individuals of MENA ancestry remain profoundly underrepresented in genomic research, comprising less than 1% of samples in major datasets (Popejoy & Fullerton, 2016). This underrepresentation reinforces calls for expanding genomic reference panels and ancestry-specific GWAS efforts to include MENA populations, thereby improving precision dementia risk prediction (Sirugo et al., 2019). Although MENA immigrants carried fewer AD risk SNPs, they reported higher rates of parental and sibling (collectively ‘family’) hypertension and diabetes diagnoses compared to the European populations. Family history of vascular and metabolic disease is a well-established marker of intergenerational cardiometabolic vulnerability in both Western (Moonesinghe et al., 2019) and MENA (Mezhal et al., 2025) populations, reflecting both shared genetics and early-life/cultural environments. These familial patterns may amplify dementia risk through lifelong exposure to vascular dysfunction, chronic inflammation, and impaired glucose regulation, all of which are potent risk factors of dementia susceptibility. This suggests that family history relevant to dementia risk may extend beyond the traditional focus on family history of memory disorders to include family history on metabolic health as well. Our interpretation is supported by clinical evidence from the MENA region. A recent UAE-based case control study identified cerebrovascular disease as the strongest independent predictor of dementia, conferring more than a sixfold increase in odds (Dhillon & Fazal, 2025b). Recognizing this broader familial risk pattern may improve personalized risk assessment and early identification of MENA individuals who are predisposed to vascular and metabolic stress decades before dementia onset.

MENA immigrants in our study were more likely to reside in neighborhoods characterized by higher material deprivation, greater exposure to air pollutants, and environments with less greenspace. Long term exposure to air pollutants impact the vascular system by promoting inflammation, oxidative stress, endothelial dysfunction, and can ultimately lead to the development of high blood pressure and atherosclerosis (Brook et al., 2010). Studies have linked chronic exposure to air pollution as a risk factor for cardiovascular disease and cognitive decline. A large longitudinal cohort study found that long-term exposure to PM_2.5_ and NO_x_ increased dementia risk by as much as 50%, with stroke accounting for nearly half of PM_2.5_ related dementia cases (Grande et al., 2020). Notably, the majority of MENA individuals (83%) migrated on or before the year 2000, leaving ample time for cumulative exposure to environmental pollutants to take place, in addition to any unmeasured exposures experienced in their countries of origin. In addition to environmental exposures, MENA immigrants experienced, on average, higher socioeconomic deprivation compared to all British, German, and Indian comparison groups. Socioeconomic disadvantage is closely linked with food insecurity, crowded housing, reduced access to health promoting activities such as exercise, and overall with chronic stress (Schultz et al., 2018). This context of disadvantage is compounded by the demographic profile of our cohort; MENA immigrants had the highest mean age at immigration, with over 80% immigrating after age 12. A later age of migration may suggest a longer period of exposure to pre-migration stressors in their countries of origin, such as political instability, armed conflict, and collective trauma, which has been identified as a significant but often unmeasured contributor to dementia incidence in MENA populations (GBD 2019 North Africa and the Middle East Neurology Collaborators, 2024). Chronic psychosocial stress stemming from these early-life exposures as well as financial strain, social instability, and migration-related challenges can have profound effects on cardiometabolic (Osborne et al., 2020; Vaccarino & Bremner, 2024) and brain health (McEwen, 2017). Such chronic stressors engage neuroendocrine pathways that elevate cortisol, impair endothelial function, promote central adiposity, and disrupt hippocampal plasticity, all of which are biological processes that accelerate metabolic aging and increase vulnerability to ADRDs (McEwen & Morrison, 2013). Chronic social stress has also been shown to upregulate pro-inflammatory cytokine activity and alter innate immune gene expression, further compounding vascular and neuroinflammatory risk (Irwin & Cole, 2011). Consistent with this framework, MENA immigrants in our study reported higher levels of loneliness and poorer self-rated health compared to European populations. This aligns with research comparing U.S.-born and immigrant Arab Americans, which found that immigrants reported higher levels of loneliness, and that loneliness was associated with poorer health (Ajrouch, 2008). Loneliness is increasingly recognized as a biological stressor that upregulates inflammatory signaling, accelerates vascular aging, and predicts faster cognitive decline (Cacioppo et al., 2015). Further network size and positive relationship quality appear to have significant effects on cognitive health for MENA older adults (Ajrouch et al., 2026). When compounded with environmental adversity and high metabolic burden, these psychosocial stressors likely exert synergistic effects that amplify brain vulnerability to ADRDs in MENA populations.

Lifestyle behaviors further differentiated MENA participants. They reported a diet that was lower in quality, poorer sleep, lower physical activity, and higher smoking prevalence relative to European immigrants. The consistently lower diet quality scores observed in the MENA group compared to the European groups across both sexes, indicate broad overconsumption of refined grains and underconsumption of whole grains, fish, dairy, and unsaturated fats. Systematic reviews and meta-analyses consistently show that higher whole-grain/fiber intake and lower refined-grain consumption are associated with reduced risk of type 2 diabetes, cardiovascular disease, and all-cause mortality (Aune et al., 2016). A narrative review corroborated these trends specifically in Middle Eastern populations, while also pointing out the need for well designed studies in this area (Al-Khudairy et al., 2013). Recent large-scale data from India demonstrate that population-wide reliance on refined carbohydrates drives profound glycemic responses and risk of type 2 diabetes (Anjana et al., 2025). The authors demonstrated through statistical modeling that replacing just 5% of daily calories from carbohydrates with that of protein from plants, dairy, egg, or fish was associated with lower risk of newly diagnosed type 2 diabetes and prediabetes (Anjana et al., 2025). Although MENA dietary patterns differ culturally from those in India, MENA immigrants had a higher intake of refined grains, higher rates of obesity, and did not differ significantly in their rates of diabetes compared to Indian populations, suggesting a metabolic risk profile at least as adverse as that observed in Indian populations. Lower intake of fish, fish oil supplements, and other unsaturated fats in the MENA group may further exacerbate vascular and neuroinflammatory pathways, given strong evidence of these compounds supporting endothelial function, reducing systemic inflammation, and protecting against cognitive decline (Bodur et al., 2025). Thus, the dietary patterns observed in the MENA cohort may contribute to dementia susceptibility both directly through diets that promote inflammation, oxidative stress, and endothelial injury and indirectly through diabetes and visceral adiposity. These findings underscore diet quality, particularly carbohydrate quality and healthy-fat intake, as a key modifiable target for dementia prevention and metabolic risk reduction in MENA communities. Importantly, diet is highly modifiable and culturally adaptable, offering a realistic opportunity for targeted dementia prevention strategies among MENA immigrants.

Dietary patterns were consistent with higher rates of obesity and diabetes in MENA populations compared to Europeans. MENA females were 4-6x and MENA males were 2-4x more likely to have diabetes compared to European populations, but not Indian populations. Increased risk of diabetes in MENA immigrants is in line with previous research comparing immigrants from MENA in the United States to US-born non-Hispanic white individuals (Kindratt et al., 2024). The biological consequences of obesity and diabetes not only depend on the quantity of adipose tissue, but also on its architecture, specifically having a propensity toward adipocyte hypertrophy rather than hyperplasia. Hyperplastic expansion, i.e., producing a greater number of smaller adipocytes, is considered a metabolically healthy pattern of fat storage. In contrast, hypertrophic fat expansion, also known as the “sick fat cell”, is characterized by fewer but much larger adipocytes (Bays, 2011). When adipocytes expand, they are pushed farther away from capillaries causing hypoxia. This hypoxia induces the secretion of proinflammatory cytokines that facilitate angiogenesis as a compensatory mechanism to restore oxygen supply (Andrei et al., 2017). However, if this expansion is too rapid or chronic, the persistent hypoxic and inflammatory milieu can lead to chronic insulin resistance, ectopic lipid deposition, inflammation, and impaired metabolic flexibility (Lundgren et al., 2007; Weyer et al., 2000). Prior work in South Asian populations demonstrates a clear bias towards hypertrophic adipose expansion (Anand et al., 2011; Chandalia et al., 2007; McLaren et al., 2024), providing a mechanistic explanation for their elevated rates of diabetes and cardiometabolic disease despite comparable or lower BMI relative to Europeans. Recent metabolomic analyses in South Asian adults demonstrate that visceral and intrahepatic fat predict incident type 2 diabetes independent of BMI, underscoring the limitations of BMI-based risk stratification in non-European populations (Gadgil et al., 2024). Although comparison studies of adipocyte morphology in MENA populations are limited, a recent study in a Tehran abdominal surgery cohort of 80 adults found that adipocyte size, but not adiposity, correlated with markers of endoplasmic reticulum stress, inflammation, and insulin resistance (Pourdashti et al., 2023). Importantly, the Global Burden of Disease study identifies the MENA super-region as having the highest age-standardized diabetes prevalence worldwide, far exceeding European estimates and mirroring the pattern seen in dementia projections (GBD 2021 Diabetes Collaborators, 2023). If MENA individuals, like South Asians, have a limited capacity for adipocyte hyperplasia, then excess caloric load may be preferentially routed into enlarged, dysfunctional adipocytes and ectopic depots (i.e., hepatic or visceral fat), accelerating metabolic deterioration at lower BMI thresholds. Biological similarities in fat storage between MENA and Indian populations is plausible given we found no difference between MENA and Indian populations in diabetes rates and body fat percentages in our study. This interpretation points to possible metabolic rigidity in MENA individuals, offering a cellular mechanism linking the elevated obesity and diabetes rates in these populations to heightened vascular and neurological risk.

Rates of obesity and diabetes were especially pronounced in MENA females. Emerging global evidence suggests that people from MENA, particularly females, are disproportionately affected by the consequences of low physical activity (Wu et al., 2025; Xu et al., 2022). Here, we found individuals from MENA had the lowest physical activity. GBD analysis show that low physical activity contributes to a larger share of diabetes and cardiovascular diseases disability adjusted life years (DALYs) in females compared to males, in addition to the MENA region already having a greater low physical activity-attributable disease burden compared to the global average (Farrokhpour et al., 2025). Given that MENA females in our study also exhibited higher comparative odds for diabetes and obesity and lower rates of physical activity, the intersection of biological susceptibility, possible cultural constraints on activity, and structural barriers to exercise likely compounds their dementia risk. For example, many females from MENA serve as primary caregivers for family members with dementia, which can increase their own stress and prevent them from focusing on their own cognitive and physical health (Mowafi et al., 2025). Together, these data point to low physical activity as a potential sex-differentiated driver of metabolic and cardiovascular risk in MENA populations, and underscores the need for gender and culturally tailored interventions that address structural barriers to physical activity. In addition to having the lowest physical activity, MENA individuals had the worst sleep scores, and MENA males in particular had the highest frequency of smoking. These adverse lifestyle behaviors may account for the higher rates of cardiovascular related conditions including coronary artery disease, hypertension, and hypercholesterolemia seen in MENA participants compared to the European populations. While total blood cholesterol levels were lower in MENA compared to the European populations, this may be due to the higher rates of participants being on cholesterol medication in MENA and India (23-38%) compared to the British and German (11-12%) populations. Despite total blood cholesterol levels being lower, particularly atherogenic particles including Lipoprotein(a) and serum triglyceride levels were significantly elevated in MENA compared to the European populations. Independent of LDL-c, elevated lipoprotein(a) adds an additional layer of risk by carrying oxidized phospholipids that accelerate plaque formation, impair fibrinolysis, and increase thrombotic potential (Gilliland et al., 2023). Elevated Lp(a) concentrations are also associated with large-artery ischemic stroke (Pan et al., 2019) and with greater white-matter hyperintensity burden indicative of small-vessel disease (Caruso et al., 2025), linking lipid abnormalities to chronic cerebral hypoperfusion, microinfarcts, and neurovascular injury.

Neuroimaging results were broadly concordant with these clinical and behavioral profiles. MENA and Indian participants combined showed reduced hippocampal volume relative to European participants despite being younger on average, suggesting an early or accelerated trajectory of hippocampal vulnerability in these groups. Reduced hippocampal volume is a well-established marker of AD as well as cardiometabolic and vascular injury and has been linked to obesity (Ambikairajah et al., 2020), insulin resistance (Frangou et al., 2022; Rasgon et al., 2011), hypertension, and chronic inflammation (Finger et al., 2022), all of which were elevated in participants from MENA. In our dataset, C-reactive protein (CRP) levels were higher in MENA participants across both sexes compared to European groups (**Tables S20-S21**), indicating a pattern of chronic low-grade systemic inflammation consistent with this elevated metabolic burden. Thus, the structural differences observed here are consistent with the disproportionate metabolic burden identified in the full sample and align with prior evidence that metabolic dysregulation may accelerate limbic system atrophy well before clinical symptoms emerge. We also observed lower global mean diffusivity (MD) in MENA and Indian participants relative to Europeans, an initially counterintuitive finding given their higher vascular and metabolic burden. However, MENA and Indian individuals were, on average, approximately two years younger than British and German participants in the imaging subset. Mean diffusivity has been shown to increase with age, reflecting cumulative microstructural degradation including myelin breakdown and expansion of extracellular space that reduces barriers to water diffusion (Bosch et al., 2012; Lawrence et al., 2021; Lebel et al., 2012). Thus, the higher MD observed in British and German participants may reflect relatively later-stage microstructural alterations that occur during normal aging. Importantly, the existence of reduced hippocampal volume suggests that distinct neurobiological processes may be unfolding on different timelines, with limbic atrophy potentially emerging earlier than widespread white matter microstructural degeneration. This dissociation may indicate that metabolic and inflammatory stressors may differentially impact gray and white matter compartments and may do so before classical diffusion signatures of aging become apparent. Collectively, these imaging findings underscore the possibility that cardiometabolic, environmental, and psychosocial exposures may imprint on brain structure through heterogeneous pathways across ethnic groups and highlight the need for ancestry-informed normative models to improve interpretation of diffusion and volumetric biomarkers in diverse populations.

Standardized geographic definitions of the Middle East and North Africa are inconsistent across international health and economic organizations and have shifted over time, with the GBD’s own definition changing since its inception. Nonetheless, to assess the robustness of our results, we performed a sensitivity analysis restricting the MENA group to the 21 countries (N=1735, **Tables S51-S73**) defined by the GBD North Africa and Middle East “super-region” (GBD 2021 Risk Factors Collaborators, 2024). While the broad categories of structural and behavioral risk factors remained significantly more adverse in the MENA population than in European and Indian comparison groups, including the highest levels of material deprivation, pollution exposure, as well as having unfavorable levels of physical activity and sleep, this more refined grouping revealed important nuances in diet quality. Overall diet quality in the GBD-restricted subset remained significantly lower in MENA immigrants compared to German and Indian populations, while no significant difference was observed relative to the British group. These findings are initially counterintuitive given that the health-promoting Mediterranean diet also has origins in the Levant (Radd-Vagenas et al., 2017). However, despite lower overall scores, GBD-defined MENA immigrants exhibited significantly higher scores for fruit and vegetable intake compared to all other groups in both males and females (**Tables S66-S67**). This may suggest that while certain core strengths of the traditional Mediterranean diet are preserved, they are increasingly offset by a profound overconsumption of refined grains and sugary foods. This dietary signature may reflect a dual burden: the gradual decline in traditional practices observed within the MENA region and culture due to rapid urbanization and globalization (Faris et al., 2025; Mendez et al., 2004), further compounded by the environmental stressors and convenience-driven food landscapes encountered post-migration to Western countries. The refined grouping also revealed important shifts in genetic and metabolic comparisons. In the broad categorization of MENA, Indian immigrants did not differ significantly from MENA in the frequency of ApoE4, consistent with previous literature identifying a north-to-south gradient in the continent of Asia, where ApoE4 is lowest in South Asia and the Mediterranean (Belloy et al., 2019). However, compared to the GBD-defined MENA group, Indian immigrants now carry a significantly higher MAF for the ApoE4 allele. While the absolute difference in frequency is modest (1%), its statistical significance indicates that MENA ApoE4 frequency may be even lower than previously defined. Furthermore, the most prominent metabolic shift in this subset was the emergence of a higher diabetic and glycemic burden in Indian immigrants compared to the MENA group. These results may reinforce the extreme metabolic rigidity and adipocyte hypertrophy characteristic of the South Asian phenotype and suggest that while both groups may face metabolic inflexibility, the MENA population’s risk may be uniquely shaped by a broader convergence of obesity and the study’s most adverse structural and environmental exposures.

Finally, this study should be interpreted in the context of several limitations. UKB’s healthy volunteer bias may underestimate disease burden across all groups (Fry et al., 2017). Healthy bias exists even within the imaging subset compared to the nonimaging subset, so underestimations of group differences may exist in our imaging subsample (Lyall et al., 2022). Age and sex mismatch was also present in our dataset, reflecting demographic characteristics inherent to the UK Biobank immigrant populations rather than study selection. To address this, we conducted sex-stratified age matched analyses in our supplementary work, which demonstrated broadly similar patterns in all factors assessed in our study (**Tables S30-S50**). Although these steps help reduce confounding, residual imbalance affecting comparisons may still exist given the underlying composition of the cohort. Moreover, migration related variables such as duration of residence, early life adversity, and pre-migration exposures were limited or absent. This is particularly relevant for stress-related measures, as many MENA immigrants may have experienced cumulative psychosocial stressors prior to migration, including political instability, prolonged armed conflict, mass displacement, and collective trauma, which have been shown to produce enduring physiological stress responses and long term cardiometabolic and neurocognitive consequences (Mowafi et al., 2025). Failure to account for these exposures may lead to underestimation of lifetime stress burden and its contribution to metabolic, vascular, and neurocognitive risk in MENA populations. Furthermore, the higher prevalence of non-substantive responses among MENA and Indian immigrants compared to Europeans, particularly for questions related to mental health, may suggest the presence of acculturation related stressors (i.e. stigma or language barriers) not fully accounted for in our analyses (Saechao et al., 2012). Nonetheless, this study represents an important step toward integrated, multimodal characterization of ADRD risk in MENA immigrant populations and highlights the need for future studies with age- and sex-balanced designs and more detailed assessment of migration-related and life course exposures.

Overall, our work suggests that dementia risk in MENA populations may be best understood through a multidimensional framework that integrates metabolic, vascular, environmental, and psychosocial vulnerability, rather than through traditional single-factor models. This multidimensional risk landscape underscores the need for ancestry and context-specific dementia models and highlights opportunities for targeted prevention in a historically underrepresented population.

## Data Availability

UK Biobank data available here: https://www.ukbiobank.ac.uk/

https://brainescience.shinyapps.io/adrd_mena_ukb/

## Supplementary Materials

Supplementary materials can be found at: https://brainescience.shinyapps.io/adrd_mena_ukb/

## Acknowledgements

This work was supported by the Alzheimer’s Association grant AARG-23-1150420, NIH grants T32AG058507, R01AG087513, and S10OD032285, R01AG081768 as well as funding by the Wellcome LEAP’s CARE initiative. It was completed under UK Biobank Resource under application number 11559.

## Footnotes

1 In the Global Burden of Disease (GBD) framework, a “super-region” is the highest level of geographic aggregation below the global level. The GBD study classifies the world’s 204 countries and territories into regions that are made up of countries and territories that are geographically close and epidemiologically similar, and regions are grouped into super-regions on the basis of cause of death patterns (GBD 2021 Risk Factors Collaborators, 2024).

